# AI regulation in healthcare around the world: what is the status quo?

**DOI:** 10.1101/2025.01.25.25321061

**Authors:** Felix Busch, Raym Geis, Yuan-Cheng Wang, Jakob Nikolas Kather, Noor Al Khori, Marcus R Makowski, Israel K Kolawole, Daniel Truhn, Warren Clements, Stephen Gilbert, Lisa C Adams, Esteban Ortiz-Prado, Keno K Bressem

**Author notes:** Correspondence: Felix Busch, MD; Mail; Address: Klinikum rechts der Isar, TUM University Hospital, Ismaninger Str. 22, 81675 Munich, Germany. Contributed equally (Esteban Ortiz-Prado, Keno K Bressem). **Funding** European Union.

## Abstract

The rapid adoption of artificial intelligence (AI) raises challenges related to ethics, safety, equity, and governance that require robust regulatory frameworks. In most jurisdictions, AI-driven medical devices are already covered by existing medical device frameworks, although new AI-specific legislation may be required to address the challenges posed by recent advancements. This expert review focuses on frameworks and legislation explicitly tailored to AI, synthesizing research literature, government and intergovernmental framework programs, and online media coverage to provide an up-to-date assessment of global AI-specific regulation or strategies in healthcare as of December 2024. Our findings show that only 15.2% (n=30/197) of countries or territories have enacted legally binding AI-specific legislation, including the 27 member states of the European Union (EU) following the adoption of the EU AI Act. A further 9.1% (n=18/197) have drafted legislation, and 28.4% (n=56/197) have issued non-binding guidelines. Notably, 47.2% (n=93/197) of countries or territories do not have an AI-specific framework or legislation in place. Furthermore, our results highlight disparities between the Global North and South, with 60.3% (n=82/136) of Global South countries or territories lacking frameworks or legislation, compared to 18% (n=11/61) in the Global North. In conclusion, our work provides an overview of the status quo of AI regulation around the world, highlights disparities in the adoption of frameworks and legislation, and calls for the need for intergovernmental and regional cooperation.

## Introduction

Artificial intelligence (AI) is increasingly being integrated into various industries worldwide, including healthcare, with a recent analysis estimating that AI could contribute up to $15.7 trillion to the global economy by 2030, driving gross domestic product increases of up to 26% in China or 14.5% in North America.^1^ In healthcare, AI promises to improve diagnostic accuracy, treatment planning, and resource management. However, global organizations such as the World Health Organization emphasize the urgent need for safeguards to ensure data security, protect patients, and promote equitable access.^2,3^ Although 67 countries have introduced national AI strategies between 2017 and 2023, many still rely on existing data privacy or medical device frameworks rather than AI-specific legally binding legislation, potentially resulting in a patchwork of oversight that could ultimately put patients at risk.^4^

Meanwhile, several intergovernmental initiatives have issued AI-specific guidelines. In March 2024, the United Nations (UN) General Assembly adopted its first resolution on AI, promoting ‘safe, secure, and trustworthy’ AI systems and urging nations to protect human rights and monitor potential risks.^5^ In September 2024, the newly created UN High-level Advisory Body on AI released its *Governing AI for Humanity* report, which suggested that while no new global AI agency is currently on the agenda, stronger international coordination might be necessary.^6^ Other intergovernmental organizations like the Organization for Economic Cooperation and Development (OECD) published various reports on AI risks, benefits, and policy imperatives and convened an Expert Group on AI Futures to address emerging challenges.^7–9^ In addition, the *G7 Hiroshima Process on Generative AI* and the *G20 AI Principles* advanced voluntary principles for AI oversight, emphasizing transparency, accountability, and data privacy.^10,11^ However, these frameworks remain non-binding, offering only voluntary guidelines for the safe and ethical adoption of AI.

In contrast, the European Union (EU) enacted the first supranational legally binding framework for AI in March 2024 with the adoption of the AI Act, which applies to all 27 member states.^12^ In addition, in September 2024, the Council of Europe opened for signature the *Framework Convention on AI and Human Rights, Democracy, and the Rule of Law*, the first internationally binding treaty that requires signatories to integrate safeguards throughout the entire AI lifecycle.^13^

In this rapidly evolving landscape, the present expert review synthesizes research literature, government and intergovernmental framework programs, and online media coverage to provide an up-to-date assessment of global AI-specific regulation and strategies in healthcare as of December 2024. In general, in most jurisdictions with established regulatory structures, medical devices, including those that integrate AI, are already subject to existing medical device frameworks. Thus, the absence of dedicated legislation does not necessarily mean AI-powered systems are unregulated. In contrast, this review centers on emerging frameworks and legislation explicitly tailored to AI with the aim to identify regional disparities and offer insights into current national AI-specific frameworks and legislation, guiding researchers, policymakers, and other stakeholders through the complexities of AI-driven healthcare governance. The subsequent sections are organized by regions or countries, depending on the specific AI frameworks or legislative measures implemented at the regional or national level.

### Search strategy and selection criteria

References for this review were identified through searches on PubMed and Google Scholar across all 197 countries and territories, using the name of each country or territory as the primary search term, followed by “artificial intelligence” combined with “strategy,” “legislation,” “regulation,” “law,” or “framework,” without time restrictions as of December 2024. Articles were also identified through searches of the authors’ own files. In addition, the authors have reviewed government and intergovernmental framework programs, as well as media reports, to determine the regulatory status of each country or territory. The final reference list has been compiled on the basis of originality and relevance to the broad scope of this review and, in most cases, refers directly to the English version of the relevant frameworks or legislation.

### European Union

The EU adopted the AI Act in March 2024, a horizontal regulatory framework for AI applying to various sectors, including healthcare, which has been binding law in all 27 member states since August 2024.^12^ The AI Act aims to protect health, safety, and fundamental rights while promoting trustworthy AI across industries and categorizes AI systems into different risk levels, each with specific regulatory requirements.^14^ Prohibited AI practices, such as manipulative or exploitative AI and facial recognition, are banned unless used in lawful medical contexts like psychological treatments. High-risk AI systems, including medical devices under the EU *Medical Device Regulation* (MDR), must now meet new standards regarding fairness, bias control, equity, transparency, and training data.^15^ General-purpose AI models, like large language models, face additional obligations if they present systemic risks, eg, if high computational power was used in their training, while minimal-risk systems, like spam filters, are subject to voluntary codes of conduct. Notably, the Act’s extraterritorial reach ensures that AI systems affecting the EU market, regardless of where they are developed or hosted, are subject to its regulations.

A key feature of the Act is its harmonization with existing EU legislation, particularly in the healthcare sector. This harmonization aims to simplify the regulatory landscape, as AI systems already subject to the MDR or *In-Vitro-Diagnostic Medical Device Regulation* (IVDR) ideally would not have to meet separate AI Act requirements and comply with the standards set by both, avoiding the need for double certification.^15–17^ However, it is anticipated that medical AI systems subject to the MDR or IVDR will still need to address both frameworks and coordinate their compliance efforts through designated notified bodies to ensure full regulatory alignment.^17^

The Act’s enforcement timeline is phased, beginning with voluntary compliance in August 2024.^18^ Key deadlines include February 2025, when AI systems posing unacceptable risks must cease operations, and August 2027, when enforcement begins for medical devices Class IIa and higher that are not subject to biometric considerations. Non-EU member states within the European Free Trade Association are expected to regulate AI in accordance with the EU AI Act (Iceland, Norway, and Liechtenstein) or to align their own proposed legislation with the Act (Switzerland). This is similar for the current EU accession candidates, including Albania, Bosnia and Herzegovina, Georgia, Moldova, Montenegro, North Macedonia, Serbia, and Ukraine.

### United Kingdom

The United Kingdom (UK) does not currently have specific legislation targeting AI, but existing regulations like the UK MDR cover software and autonomous decision-making systems, including AI, even though they were not explicitly designed for recent advancements in machine learning, deep learning, or generative AI. As outlined in a white paper titled *AI regulation: a pro-innovation approach* from the Department for Science, Innovation and Technology and Office for AI in August 2023, the UK’s approach to AI regulation prioritizes a flexible, principles-based, and non-statutory framework interpreted by sector-specific regulators rather than introducing horizontal legislation like the EU AI Act.^19^ In the healthcare sector, the Medicines and Healthcare products Regulatory Agency (MHRA) announced the *Software and AI as a Medical Device (SaMD/AIaMD) Change Programme* in 2021 to ensure the UK MDR is fit for purpose for AIaMD.^20^ At the current stage, this non-binding framework provides guidance on how to address challenges like transparency, risk management, and lifecycle validation for AIaMD and aims to complement the UK MDR. More recently, the government committed to regulating AI in July 2024.^21^ Since then, however, the new administration has not proposed any specific legislation on AI.

### United States

In the United States (US), while there is no horizontal federal legislation explicitly governing AI, existing laws such as the *Federal Food, Drug, and Cosmetic Act*, as amended by the *1976 Medical Device Amendments* and subsequent updates, regulate software, including autonomous systems and AI-based decision-making tools.^22^ These regulations are enforced by the Food and Drug Administration (FDA), which has approved 1,016 AI-and machine-learning-enabled medical devices as of December 2024.^23^ The FDA also addresses AI-specific challenges through initiatives such as the *21st Century Cures Act*, which exempts certain clinical decision support systems from regulation if they meet specific criteria, and detailed guidance on AI and machine learning software used in medical devices.^24^ Additionally, the Office of the National Coordinator for Health Information Technology Systems’ *Health Data, Technology, and Interoperability Certification Program* (HTI-1 Rule) regulates AI-based decision support systems within electronic health records, providing binding requirements for their interoperability and safety.^25^

The Biden administration’s October 2023 *Executive Order on the Safe, Secure, and Trustworthy Development and Use of AI* (AI EO) directs federal agencies to develop standards, regulations, and oversight mechanisms for AI.^26^ For healthcare, the EO guides the Department of Health and Human Services, including the FDA and the Centers for Medicare and Medicaid Services, in shaping strategies for AI adoption. This includes implementing provisions from the *Blueprint for the AI Bill of Rights* and earlier frameworks like the *National AI Initiative Act of 2020*, as well as forming the Office of the Chief AI Officer to oversee these efforts.^27,28^ While these measures address issues such as transparency, risk management, and lifecycle validation for AI-based medical products, the EO itself is not statutory law, leaving its longevity uncertain amid administrative changes in January 2025.

Furthermore, states like California have proposed their own AI-specific legislation, such as the *Safe and Secure Innovation for Frontier AI Systems Act (Senate Bill SB-1047)*, including provisions for mandatory safety incident reporting, requirements for conducting safety assessments, third-party testing of AI models to ensure compliance with established standards, and civil penalties for non-compliance.^29^ Meanwhile, federal efforts like the stalled *Algorithmic Accountability Act of 2022*, which aimed to establish assessment requirements for high-risk AI systems, underscore the ongoing debates surrounding comprehensive AI legislation.^30^ Finally, the FDA has issued additional AI-specific guidance, such as its *Marketing Submission Recommendations for Predetermined Change Control Plans*, which focuses on managing updates in AI-enabled medical devices.^31^

### Canada

In Canada, the regulation of AI in healthcare remains a work in progress. At the federal level, *the AI and Data Act* (AIDA) was introduced in June 2022 as part of Bill C-27, the *Digital Charter Implementation Act*.^32^ AIDA proposes a risk-based framework for regulating AI systems, particularly high-impact applications, including those in healthcare, by requiring transparency and harm mitigation measures. However, as of December 2024, AIDA remains under parliamentary review and has yet to be enacted into law.

Furthermore, the 2023 federal *Guide on the Use of Generative AI* provides principles and best practices for responsible AI adoption within Canadian government operations.^33^ It emphasizes ethical considerations, transparency, and accountability in deploying generative AI systems to support public services while mitigating risks.

Above all, in 2021, Health Canada, the FDA, and the UK MHRA jointly published the *Good Machine Learning Practice for Medical Device Development: Guiding Principles*, a framework that addresses the safety, efficacy, and transparency of AI and machine learning-enabled medical devices.^34^ In June 2024, the authorities expanded this framework by introducing the *Transparency for Machine Learning-Enabled Medical Devices: Guiding Principles*, which provide for clear communication about the performance, intended use, development process, and logic of the devices.^35^ Although, again, not legally binding frameworks, these principles aim to inform stakeholders in the healthcare sector, promote trust, and encourage informed use throughout the lifecycle of AI-driven medical devices in the UK, US, and Canada.

### Mexico

In recent years, several digital health legislative initiatives have been proposed to the Mexican Senate, but none have been passed.^36^ As of December 2024, the regulation of AI in healthcare remains under the jurisdiction of existing legal frameworks, including the *General Health Act* and the *Protection of Personal Data Held by Private Parties*.^37,38^ In February 2024, a new legislative proposal, the *Federal Law Regulating AI*, was introduced to the Mexican Senate.^39^ This proposed law aims to establish a legal framework for the development, deployment, and use of AI technologies across various sectors, including healthcare. Moreover, it proposes designating the Federal Telecommunications Institute (IFT) as the competent authority on AI matters and suggests the creation of a National Commission on AI to serve as an advisory body to the IFT. The law also classifies AI systems by risk levels as unacceptable, high, low, or minimal, similar to the EU AI Act, and outlines specific obligations and testing requirements for each category. Developers or providers of AI systems would be required to obtain prior authorization from the IFT before commercializing their systems in Mexico, even if offered free of charge. However, as the legislative process involves multiple stages of review and approval, the exact timeline for the law’s adoption remains uncertain. Furthermore, as with other previously proposed digital health legislation, it is still possible that the law will not pass the Senate.

### Central America and the Caribbean

As of December 2024, countries in Central America and the sovereign states in the Caribbean have not implemented binding legislation specifically regulating AI. Instead, some countries have introduced national AI strategies or are participating in non-binding regional initiatives.

In Central America, Costa Rica was the first country to announce its national AI strategy, while the Dominican Republic was the first country in the Caribbean. Both frameworks focus on principles of ethical use, privacy, and societal benefit, serving as strategic guidance rather than legally enforceable regulations.^40,41^

Regional initiatives, such as the *UNESCO Caribbean AI Policy Roadmap*, provide a foundation for promoting innovation and establishing oversight for AI but do not constitute binding legislation.^42^ Similarly, in August 2024, 17 Latin American and Caribbean countries adopted the *Cartagena de Indias Declaration on AI*.^43^ However, the declaration itself does not establish enforceable legal mechanisms but rather serves as a collaborative guideline for future policy development.

### South America

In South America, no country has enacted AI-specific legislation. However, six countries, namely Brazil, Peru, Chile, Argentina, Colombia, and Ecuador, have drafted legal frameworks for AI.

Brazil’s national AI strategy, based on the OECD AI Principles, was launched in 2021.^44^ In December 2024, the Brazilian Senate approved an AI Framework (*Bill No. 2338/2023*), which was first proposed in May 2023.^45^ This bill establishes a horizontal regulatory framework for AI, adopting a risk-based approach similar to the EU AI Act and implementing a dedicated regulatory authority. Until this framework is implemented, medical AI applications (classified as SaMD) are regulated by the National Health Surveillance Agency (ANVISA) under the existing ANVISA regulatory framework.^46^

In Peru, an updated draft of its AI governance legislation (*Law No. 31814*) was introduced in November 2024, building on the original proposal from May 2023.^47^ The law adopts a risk-based framework for classifying AI systems and emphasizes developer accountability for ensuring safety, explainability, and reliability. Earlier, in 2020, Peru had enacted *Emergency Decree 007-2020*, which provided foundational guidelines for secure digital interactions, including principles and structures for oversight, monitoring, and cybersecurity.^48^ So far, this decree encompasses AI within its broader regulatory scope, including AI-driven healthcare applications.

Chile issued its national AI policy in 2020, after which *Bill No. 16821-19* was developed and submitted to the Chamber of Deputies in May 2024.^49,50^ The bill adopts a risk-based classification similar to the EU’s AI Act and imposes obligations on high-risk applications, such as data governance, transparency, and human oversight. It also establishes the AI Technical Advisory Council, and grants enforcement authority to the Personal Data Protection Agency. As of December 2024, the bill is still under deliberation, and AI applications in healthcare fall under Chile’s MDR framework.^51^

In 2021, Argentina began drafting a national AI plan as part of its *Innovative Argentina 2030 Plan* and *2030 Digital Agenda*.^52^ In June 2024, the Argentine Congress introduced *Bill 3003-D-2024*, which aims to establish a legal framework for the responsible use of AI.^53^ As in other South American countries, the proposed legislation is inspired by the EU’s AI Act and takes a risk-based approach to defining the obligations of AI providers and users. To date, AI-driven SaMD is regulated under the Argentine MDR.^54^

Colombia is also in the process of passing binding AI-specific legislation. In November 2023, the Senate passed *Law 059/23*, which establishes guidelines for the development, use, and regulation of AI as part of the *National Digital Strategy 2023-2026*.^55,56^ The law emphasizes principles such as human oversight of AI, safety, cooperation, and the public good. It also calls for the establishment of a Commission on Data Processing and AI Development to draft technical regulations, advise Congress, and promote AI-driven innovation. As of December 2024, the law is still under review, and SaMDs are regulated under the National Institute for Food and Drug Surveillance’s MDR.

Ecuador’s efforts to adopt AI legislation resulted in the proposal of a draft *Organic Law for the Regulation and Promotion of AI* in July 2024, which establishes a risk-based framework for the safe and ethical use of AI while promoting innovation across sectors.^57^ This follows earlier initiatives, such as the *2022 Digital Transformation Agenda* and the establishment of an AI Committee under the *Organic Law for Digital and Audiovisual Transformation* in 2023.^58,59^ Until the law is enacted, SaMDs are governed by the National Agency for Regulation’s medical device framework.

The last country in South America to draft a national AI strategy for public consultation was Uruguay in October 2024.^60^ Other countries, including Venezuela, Paraguay, and Bolivia, have yet to develop national frameworks or introduce AI-specific legislation.

### China

Over the past few years, China’s regulatory framework for governing AI applications in healthcare has developed steadily, mainly under the direction of the National Medical Products Administration and the National Health Commission. Starting in July 2021, the *Guidelines for the Classification and Definition of AI Medical Software Products* were issued to define management attributes and risk-based classifications for AI-based medical software.^61^ Concurrently, the *Guidelines for the Naming of General Names for Medical Software* standardized naming conventions by requiring a core term and two descriptive feature terms for product clarity and consistency.^62^

In March 2022, the regulatory landscape expanded with the *Guidelines for the Registration and Review of AI Medical Devices*, which standardized submission requirements and technical review processes, alongside the *Guidelines for the Registration and Review of Medical Device Software (2022 Revised Edition)* and the *Guidelines for the Registration and Review of Medical Device Cybersecurity (2022 Revised Edition)*, guiding the technical evaluation and foster cybersecurity.^63–65^ In addition to the cross-specialty guidance of AI applications in healthcare, one of the first guidelines for specific medical applications of AI was issued with the *Guidelines for the Registration Review of Pulmonary Nodule CT Image Assisted Detection Software* in May 2022, followed by the *Guidelines for the Registration and Review of Diabetic Retinopathy Fundus Image Aided Diagnosis Software* in July 2022.^66,67^

In July 2023, China introduced the *Interim Measures for the Administration of Generative AI Services* to regulate generative AI applications across industries, emphasizing ethical governance, transparency, and risk mitigation.^68^ In addition, Chinese legal scholars proposed a draft *AI Law* in March 2024, which aims to provide a horizontal legal framework for AI developed and deployed in China.^69^ It includes provisions on AI-related risks, intellectual property, data security, and bias mitigation across industries and may complement or replace some of the introduced sector-specific guidelines.

Most recently, the National Medical Products Administration released the *Medical Device Administration Law* for public consultation on August 28, 2024.^70^ This draft law presents the first overarching regulatory framework in China for the entire life cycle of medical devices, including research and development (R&D), manufacturing, distribution, and use. It remains to be seen whether this law will also apply to AI-based medical devices, potentially replacing previous administrative regulations with limited legal authority.

### Japan

In Japan, several non-binding general guidelines for the development and use of AI have been published in recent years. Notable among these are the *AI R&D Guidelines* (2017), the *AI Utilization Guidelines* (2019), and the *Governance Guidelines for Implementation of AI Principles Version 1.1* (2022).^71–73^ These have recently been updated and consolidated into *the AI Guidelines for Business Version 1.0*, which was released in April 2024.^74^ However, all of these guidelines are intended to serve as ‘soft law’ without legally binding responsibility and do not explicitly target AI applications in healthcare.

At the same time, as part of the G7 Hiroshima AI Process, Japan contributed to the development of the *Hiroshima Process International Guiding Principles for Organizations Developing Advanced AI Systems*.^10^ These principles aim to promote safe, secure, and trustworthy AI worldwide and provide guidance for organizations developing and using advanced AI systems, including foundational models and generative AI systems.

An AI Strategy Council, established in May 2023, has also produced a *Tentative Summary of AI Issues*, highlighting the need to revise existing guidelines in light of the proliferation of generative AI.^75^

In addition, in February 2024, a working group proposed basic concepts for a *Basic Law for the Promotion of Responsible AI*, which would introduce binding legal requirements for certain generative AI foundational models.^76^ However, as of December 2024, there is no publicly available draft. If adopted, this framework would shift Japan’s previous reliance on soft law to a legally enforceable model, at least for advanced AI models, including regulation of their use in healthcare.

### Korea

In 2019, the South Korean government released its national AI strategy to foster AI adoption, including investments in R&D, workforce training, and ethical AI practices to support responsible innovation, followed by several national and international initiatives, such as the *Digital New Deal* and the *Seoul Declaration for Safe, Innovative and Inclusive AI*.^77–79^

While multiple AI-specific bills were introduced during the 21st National Assembly, these proposals failed to pass before the Assembly’s term ended in May 2024.^80^ However, as a legally binding effort to regulate AI-driven decision-making, the *Personal Information Protection Act* was amended in March 2023. These amendments give individuals the right to opt out of AI-driven decisions that affect their rights or to request explanations for such decisions.^81^

Most recently, the 22nd National Assembly passed a legal framework on the *Development of AI and Creation of Trust Base* (AI Basic Act) through the subcommittee stage in November 2024, which consolidates nineteen previous AI-related proposals into a single framework.^82,83^ The draft law adopts a risk-based approach, imposes transparency requirements, establishes risk management systems, and enforces penalties for non-compliance, mirroring elements of the EU AI Act. The South Korean Act now awaits approval by the Judiciary Committee and a final plenary session. If enacted, it will serve as South Korea’s first overarching legislation dedicated to AI, including the regulation of AI applications in the healthcare sector. Concurrently, the government plans to establish an AI Safety Research Institute under the Ministry of Science and Information and Communication Technology.^84^

In contrast to South Korea’s proactive approach to AI governance, North Korea does not have any publicly available formal regulations or policies governing the development or application of AI as of December 2024.

### South Asia

As of December 2024, no country in South Asia, including Afghanistan, Bangladesh, Bhutan, India, Maldives, Nepal, Pakistan, and Sri Lanka, has enacted AI-specific legislation.

In May 2023, Pakistan’s Ministry of Information Technology and Telecommunications unveiled a draft *National AI Policy* as part of its Digital Pakistan vision.^85^ Although the policy does not elaborate on specific regulatory approaches, it proposes the establishment of an AI Directorate under the National Commission for Personal Data Protection to oversee the ethical use of AI and suggests the creation of AI regulatory sandboxes for controlled implementation of AI. However, as of December 2024, the policy remains in draft form.

In India, the Ministry of Electronics and Information Technology announced plans to introduce a Digital India Act (DIA) to replace the existing Information Technology Act of 2000.^86,87^ The DIA aims to address contemporary challenges in the digital ecosystem, including the regulation of emerging technologies such as AI. However, as of December 2024, the draft has not been released for public consultation.

Other countries, such as Bangladesh, Nepal, and Sri Lanka, have issued national AI strategies and concept papers, some of which aim to establish a regulatory framework for AI, but without any concrete provisions to date.^88–90^ Most recently, the Maldivian government announced that it will develop an AI master plan in October 2024.^91^ Countries such as Bhutan and Afghanistan have not explicitly issued national policies or strategies.

### Southeast Asia

In Southeast Asia, seven Association of Southeast Asian Nations (ASEAN) member states, including Singapore, Vietnam, Thailand, Indonesia, Malaysia, Cambodia, and the Philippines, have introduced national AI strategies. At the regional level, ASEAN launched the *ASEAN Guide on AI Governance and Ethics* in February 2024, adopting a market-driven approach to balance AI’s benefits with its risks.^92^ However, none of these countries have legally binding AI legislation as of December 2024, with Thailand, the Philippines, and Vietnam being the only countries that issued draft laws specifically dedicated to AI regulation.

Singapore does not have specific laws directly regulating AI but adopts a soft-law approach through frameworks and sectoral guidelines. Key initiatives include the *Model AI Governance Framework* (2019, updated in 2020), the *AI Verify* governance toolkit, and the *National AI Strategy 2.0* (2023).^93–95^ In May 2024, a draft *Generative AI Governance Framework* was issued to address emerging AI challenges, providing updated recommendations for the secure development and adoption of AI, and is currently under public consultation.^96^

Currently, Indonesia regulates AI applications under the pre-existing *Electronic Information and Transactions Law*, which was amended to define AI as an electronic agent.^97,98^ Key principles for AI operators include prudence, system security, cost efficiency, and consumer protection, with liability typically falling on operators unless user negligence is proven. In 2020, the *National Strategy on AI 2020–2045* (Stranas KA) was introduced as a framework for ministries and stakeholders to guide AI policy and development.^99^ In December 2023, the government adopted non-legally binding key ethical guidelines (*Circular Letter No. 9 of 2023*).^100^ More recently, in September 2024, the Deputy Minister of Communication and Information announced plans to issue a ministerial regulation on AI, aiming to provide guidelines for AI use, though no draft or concrete details have been released so far.^101^

Thailand does not have any AI-specific legislation in force. However, in early 2023, two draft laws were introduced: the *Royal Decree on Business Operations that Use AI Systems*, which proposes a risk-based regulatory framework, and the *Draft Act on the Promotion and Support of AI Innovations*, designed to promote AI development while ensuring consumer protection.^102,103^ In addition, Thailand has issued non-binding *AI Ethics Guidelines* and *AI Governance Guidelines* to provide interim principles for ethical AI development and deployment.^104,105^

Similarly, Vietnam’s Ministry of Information and Communication issued a draft *Digital Technology Industry Law* for public consultation, proposing regulations for AI.^106^ The draft outlines ethical principles for AI development and use, requires labeling of AI-generated digital products to ensure they are identifiable as artificially created or manipulated, and introduces a risk-based classification of AI systems based on their impact on health, safety, rights, and critical infrastructure, with regulatory measures tailored to each risk level.

In Malaysia, the government published a *National AI Roadmap* in 2023, under which a National AI Office was launched in December 2024 to oversee policy formulation and regulatory frameworks.^107,108^ Additionally, The *National Guidelines on AI Governance and Ethics* published in September 2024 provide foundational principles for ethical AI use, although not legally binding.^109^

On March 1, 2023, the Philippines introduced the *AI Development and Regulation Act (HB7396)* in the House of Representatives.^110^ The Act aims to promote safe AI development that is aligned with ethical principles and public interest. It proposes the establishment of an AI Development Authority to oversee the licensing, certification, and regulation of AI developers and deployers, requiring mandatory registration and compliance with issued guidelines. Non-compliance would result in penalties, with the Act set to take effect 15 days after publication in the *Official Gazette*. However, there have been no updates to the law since the publication of the draft.

Other ASEAN member states, such as Brunei, are still in the process of developing non-binding codes and guidelines to govern the use of AI.^111^

### Russian Federation

The Russian Federation was one of the first countries to adopt AI-specific legislation. Beginning in 2019, Russia adopted its *National Strategy for the Development of AI (2019-2030)*, which highlights the role of AI innovation in the country while emphasizing the need for regulation to ensure safety and ethical compliance.^112^ In 2020, *Federal Law No. 123-FZ* was adopted with the aim of creating experimental legal regimes through the establishment of regulatory sandboxes for AI technologies in Moscow, allowing for the testing of AI systems in controlled environments.^113^ With the original framework set to expire in 2025, policymakers have drafted an updated law on AI that will expand on the original federal law with regulations on accountability for AI actions and intellectual property ownership of AI-generated outputs.^114^ Specific to the healthcare sector, the Ministry of Health issued standards for the certification of (AI-based) medical software in August 2021, requiring conformity assessment in compliance with Russian government standards, focusing on data security and system reliability.^115^ In addition, *Federal Law No. 258-FZ*, adopted in 2021, established a nationwide legal framework for regulatory sandboxes in the field of digital innovation, including medical activities.^116^

### Central Asia, South Caucasus, and Mongolia

Mongolia and countries in Central Asia and the South Caucasus currently lack AI-specific legislation. Kazakhstan, Central Asia’s largest economy, is focusing on advancing its AI ecosystem through national initiatives like the *Digital Transformation and Cybersecurity Concept (2023–2029)* and the *Concept for the Development of AI for 2024-2029*, including a national AI platform, and an AI Committee tasked with creating and implementing policies.^117,118^ Although an AI-specific law is planned as part of the AI development concept, no draft has been finalized as of December 2024. Notably, Kazakhstan has also introduced AI sandboxes that focus on the safe development of AI in controlled environments, similar to Uzbekistan, where a national AI strategy has also been adopted.^119,120^ On the other hand, Tajikistan and Kyrgyzstan have also adopted national AI strategies, but these do not address the regulation of AI or the establishment of AI regulatory sandboxes, while Turkmenistan has yet to adopt a national AI strategy or legislation.^121,122^

At the regional level, the Commonwealth of Independent States introduced a non-binding model AI law in April 2024 to promote uniform regulatory approaches, though its practical impact remains questionable.^123^ Similarly, the Organization of Turkic States adopted the *Garabagh Declaration* in July 2024, signaling intentions to collaborate on AI policies, but concrete details are yet to be released.^124^

In the South Caucasus, Azerbaijan and Armenia are still in the process of developing national AI strategies and lack specific regulatory frameworks.^125,126^ In contrast, Mongolia adopted an action plan for 2024-2028 in August 2024, outlining goals for regulatory and big data policy improvements, but details have yet to be published.^127^

### Middle East

The Middle East presents a dynamic yet heterogeneous landscape for the regulation of AI in healthcare, characterized by countries at different stages of developing and implementing regulatory frameworks. Advanced economies such as the United Arab Emirates (UAE), Saudi Arabia, Qatar, Turkey, and Israel are leading the region with structured approaches, although in many cases, regulatory efforts focus on non-binding guidelines rather than enforceable legislation. Most notably, Bahrain was the first country in the Middle East to adopt a stand-alone law to regulate AI in April 2024, which includes 38 articles on privacy, unlicensed activities, and the risks of AI systems, with penalties for non-compliance and the establishment of an AI judicial unit for oversight and enforcement.^128^

In the UAE, the regulatory landscape is characterized by a combination of federal initiatives and jurisdiction-specific frameworks, particularly in its financial free zones, such as the Abu Dhabi Global Market and the Dubai International Financial Centre (DIFC).^129^ At the federal level, the UAE appointed a Minister for AI in 2017, followed by issuing the *UAE National Strategy for AI 2031* in 2018 and several non-binding guidelines aimed at integrating AI across various sectors.^130^ Additionally, the UAE published the *UAE Charter for the Development and Use of AI* in June 2024, which serves as a guiding framework to protect the rights of the UAE community in the development or use of AI solutions and technologies.^131^ In healthcare, certain regulatory measures for AI are being implemented through existing legal frameworks. For example, the DIFC has amended its data protection regulations to address the use of AI in personal data processing, imposing obligations on entities operating autonomous and semi-autonomous systems and emphasizing responsible data management.^132^ However, as of December 2024, neither the UAE mainland nor the financial free zones have enacted binding, AI-specific regulations.

Saudi Arabia has not issued any AI-specific legislation and instead focuses on non-binding guidelines. In September 2023, the Saudi Data and AI Authority (SDAIA) released a draft entitled *AI Ethics Principles*, which focuses on fairness, privacy, transparency, accountability, and security.^133^ In healthcare, the Saudi Food and Drug Authority has introduced a guidance document for AI and machine learning-based medical devices, which addresses safety, data governance, and compliance.^134^ More recently, the SDAIA issued the *AI Adoption Framework* in September 2024, which provides guidelines for the responsible integration and implementation of AI technologies across sectors, emphasizing ethical principles, transparency, accountability, and alignment with Saudi Arabia’s *Vision 2030*.^135,136^

Qatar, aligning with its *National Vision 2030*, launched a national AI strategy in 2019 and established an AI Committee in 2021 to oversee its implementation.^137,138^ In 2024, Qatar’s National Cybersecurity Agency issued the non-binding *Guidelines for Secure Usage and Adoption of AI,* emphasizing responsible use, risk management, ethical principles, and stakeholder confidence, with a specific focus on generative AI and organizational compliance.^139^

Turkey, while aligned with EU regulations like MDR/IVDR under the Customs Union and not directly covered by the EU AI Act, is actively developing its own AI regulatory framework. In August 2021, the *National AI Strategy (2021-2025)* was issued, emphasizing the goal to align Turkey’s AI governance with international standards, promoting the responsible use of AI across sectors, including healthcare.^140^ Building on this strategy, a *Draft Law on AI* was submitted to the Grand National Assembly in June 2024.^141^ While the draft law outlines principles such as safety, transparency, fairness, accountability, and data protection and defines roles within the AI ecosystem, it contains limited provisions on the regulation of AI.^142^ It remains to be determined whether this law will be implemented in its current form, expanded upon, or supplemented by additional regulations to achieve a more comprehensive regulatory framework.

After publishing a white paper on the responsible development of AI in November 2022, Israel’s Ministry of Innovation, Science, and Technology of Israel, in collaboration with the Ministry of Justice, proposed its first *Policy on AI Regulation and Ethics in December* (AI Policy) in December 2023.^143^ The AI Policy emphasizes human-centric AI, equality, transparency, accountability, and system reliability, adopting a risk-based, sector-specific approach aligned with international standards like the EU AI Act. However, this policy aims at ‘soft regulation’ through the use of non-binding ethical principles, voluntary standards, and both monitored and unmonitored self-regulation.

Other countries in the Middle East, including Jordan, Lebanon, Oman, Kuwait, and Palestine, have drafted or adopted national AI strategies; however, these strategies remain primarily focused on promoting innovation rather than establishing specific legislative frameworks for AI governance.^144–148^ In contrast, countries like Yemen, Iran, Iraq, and Syria currently have no formal AI framework or policy in place.

### African Union

Within the African Union (AU), several countries have issued national AI strategies that predominantly focus on capacity building, research, and innovation rather than on the regulation of AI, with healthcare often highlighted as a priority sector. Most recently, Algeria adopted its national AI strategy in December 2024, aiming to promote AI technologies in key sectors such as agriculture, health, and industry while simultaneously working on a regulatory framework for AI.^149^ As such, Algeria follows the lead of other AU member states that published national strategies, including Morocco, Zambia, Senegal, Nigeria, South Africa, Ethiopia, and Libya in 2024, Benin in 2023, Ghana and Rwanda in 2022, Egypt in 2019, and Tunisia and Mauritius in 2018.^150–162^

Other countries within the AU, like Kenya and Tanzania, are actively developing AI strategies. In April 2024, Kenya’s Ministry of Information and Communication Technology and Digital Economy, in partnership with the German International Cooperation Society, launched the national AI strategy development process.^163^ In the past, Kenya has also proposed the *Kenya Robotics and AI Society Bill* to promote the ethical and responsible development of AI technologies, which, however, is still in the draft stage.^164^

Tanzania issued a *Policy Framework for AI in Tanzania Health Sector* in 2022 and is reportedly in the process of formulating a national AI strategy for the management of AI and the protection of the public.^165,166^

On a continental level, the AU has been proactive in advancing AI governance. Following initiatives like the AU’s *Digital Transformation Strategy for Africa (2020-2030)* and the *AI Data Policy Framework of 2022*, the AU Executive Council endorsed the *Continental AI Strategy* in July 2024, a framework to guide AI governance and development across its 55 member states.^4,167^ The strategy prioritizes harnessing AI’s benefits, building capabilities, mitigating risks, stimulating investments, and fostering cooperation. Central to the strategy is AI governance, viewed as critical for responsible AI adoption, addressing societal, ethical, and legal challenges while aligning with Africa’s development goals under the *Agenda 2063*.^168^ Key measures include implementing or updating existing legal frameworks (eg, on data protection and cybersecurity), identifying regulatory gaps, and fostering cross-border data sharing to support AI development. The strategy also advocates for independent oversight bodies, ethical principles, and collaborative efforts among stakeholders to balance innovation and risk management. Implementation is planned in phases (2025-2030), aiming to harmonize AI governance across member states to ensure inclusive, fair, and sustainable AI-driven growth. Since most national strategies do not focus on the regulation of AI, the AU’s anticipated *Continental AI Strategy* marks a critical step towards a uniform development of legislation, including the application of AI in healthcare. While national strategies aim to build infrastructure, capacity, and awareness around AI, regulatory gaps remain that the continental framework could address.

### Oceania

In Australia, the *Therapeutic Goods Act 1989*, which includes the regulation of medical devices, and the *Privacy Act 1988*, which includes ensuring the security of patient data and health records, have undergone significant reforms to adapt to the rapidly evolving AI landscape since 2021, with the most recent update in August 2024.^169–171^ The purpose of these reforms is to ensure that medical AI devices are evaluated and assessed for safety while balancing the risks of over-regulation. Notably, the concept is similar to the EU’s MDR in many aspects, following a risk-based approach. Furthermore, the Therapeutic Goods Administration’s (TGA) regulatory principles include a specific provision for transparency, which excludes companies that rely on the ‘black box’ approach to protect their technology.

In New Zealand, the *Therapeutic Products Act 2023* was introduced to regulate SaMD; however, it does not include specific provisions for AI technologies.^172^ Healthcare data in New Zealand is governed under the *Privacy Act 2020*, which provides a framework for the use and protection of patient data but similarly does not explicitly address AI.^173^ Thus, similar to much of the Oceania region, including smaller Pacific island states, there are currently no specific regulations governing the use of AI in New Zealand.

Looking ahead, the Australian Government is actively reviewing existing legislation and has initiated public and stakeholder consultation on mandatory guardrails for AI through a document titled *Safe and Responsible AI in Health Care* – *Legislation and Regulation Review* (Proposals Paper).^174^ Concurrently, the TGA has recently completed its own consultation to modernize AI regulation, aiming to streamline laws that protect patients while fostering innovation and improving healthcare outcomes.^175^ In New Zealand, the Ministry of Business, Innovation, and Employment published a cabinet paper in July 2024 detailing its strategy for AI regulation, emphasizing the importance of encouraging AI adoption through a ‘light-touch,’ balanced, and risk-based approach.^176^

### Conclusions and Outlook

Our analysis provides a global overview of the regulatory landscape for AI, with a focus on healthcare, covering the status quo of 197 countries and territories as of December 2024 (see Figure 1). As a reference for the reader, we provide an interactive online version of Figure 1, where details of the regulatory status for each country and territory can be accessed, and which we intend to update regularly.^177^

**Figure 1.**
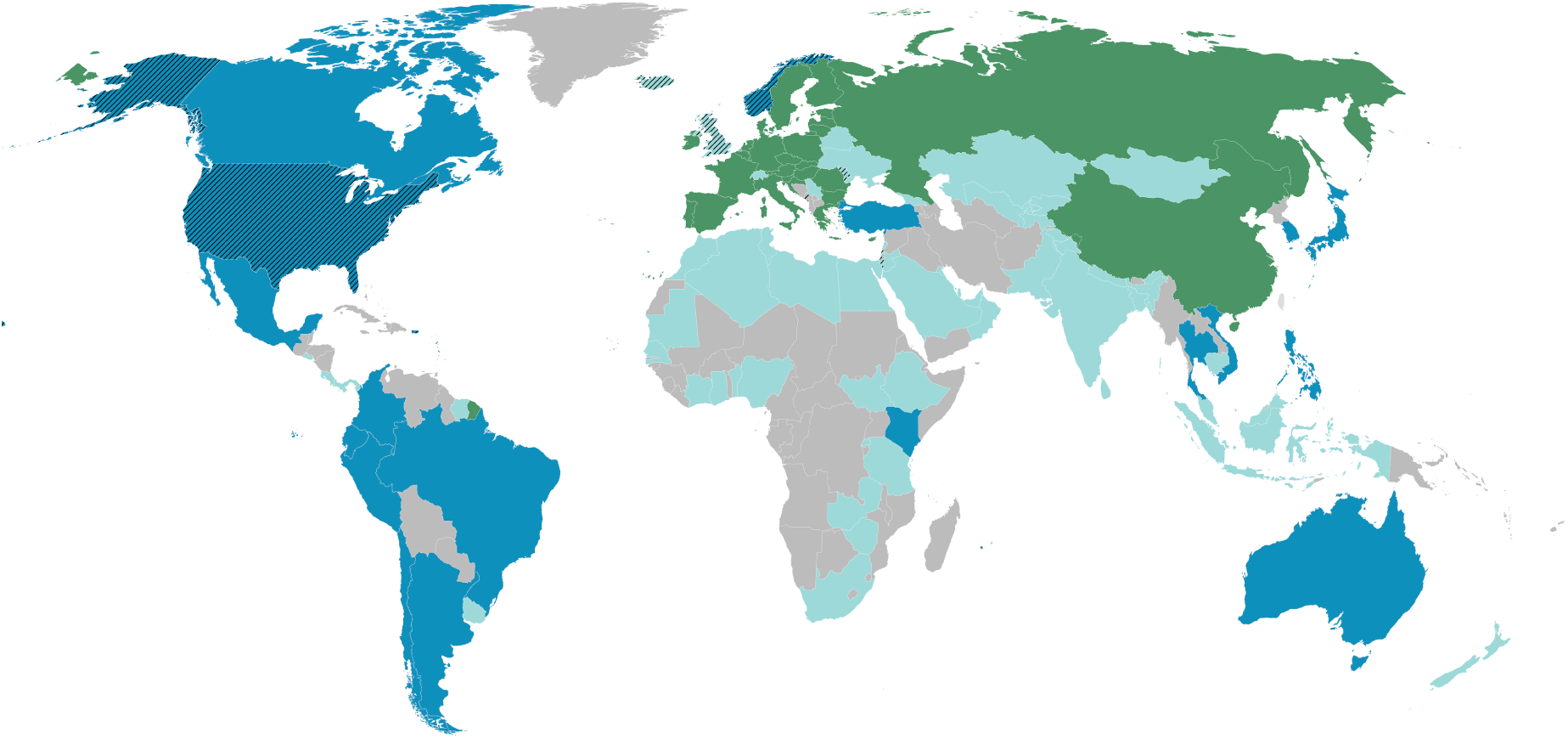
World map depicting the regulatory landscape of artificial intelligence (AI) as of December 2024. Countries or territories are color-coded based on their regulatory approach. Dark green represents countries or territories that have adopted AI-specific legislation, while dark blue indicates those that have drafted AI-specific legislation but have not yet adopted it. Turquoise represents countries or territories that have issued non-legally binding national AI guidance documents. Grey-highlighted countries or territories have neither introduced AI-specific legislation nor issued guidance. Countries or territories patterned with black diagonal lines have signed the Council of Europe’s Framework Convention on AI. We also provide an interactive online version of this map, which provides details on the status of individual countries or territories when hovered over and which the authors intend to update regularly.^177^

We found that only 15.2% (n=30/197) of countries or territories have implemented legally binding AI-specific legislation (ie, legislation explicitly tailored to AI). Notably, the 27 EU Member States account for 90% (n=27/30) of countries with legislation following the recent adoption of the EU AI Act. The remaining three countries, Bahrain, the Russian Federation, and China, have adopted different national AI legislative frameworks that reflect their national priorities and strategies.

A further 9.1% (n=18/197) of countries or territories have drafted AI-specific legislation, signaling their intention to implement legally binding frameworks, although these drafts are still under review or in the legislative pipeline. Meanwhile, 28.4% (n=56/197) have issued non-legally binding guidance documents that provide recommendations or frameworks for AI development and governance. Notably, these documents often focus on ethical principles, transparency, and fostering innovation rather than adopting AI-specific regulatory frameworks. Finally, 47.2% (n=93/197) of countries or territories have yet to introduce any form of AI-specific legislation or guidance. This lack of regulatory guidance highlights disparities in global readiness for AI governance, particularly when comparing the Global North and South. Of the 136 countries or territories classified as Global South according to the Finance Center for South-South Cooperation, 60.3% (n=82/136) had no national framework or AI-specific legislation, compared to 18% (n=11/61) in the Global North.^178^ Meanwhile, 30.9% (n=42/136) in the Global South had national AI strategies, compared to 23% (n=14/61) in the Global North. Furthermore, 7.4% (n=10/136) of countries or territories in the Global South had drafted AI-specific legislation, compared with 13.1% (n=8/61) in the Global North, and only 1.5% (n=2/136) in the Global South had enacted such legislation, compared with 45.9% (n=28/61) in the Global North.

Most importantly, the lack of such targeted legislation does not necessarily mean that AI-based medical devices are unregulated. Rather, these technologies are subject to the same safety, quality, and efficacy standards that apply to all medical devices in most countries and territories. Many in the medical device industry view an approach that focuses on updating existing regulations, such as the EU’s MDR and *General Data Protection Regulation*, as a more effective alternative to enacting entirely new horizontal legislation.^179,180^ For example, although the EU’s AI Act will be closely aligned with the MDR/IVDR, there is no evidence at present to suggest that providing updated guidance and standards under the current medical device framework will be any less effective in regulating AI-driven healthcare solutions.

However, for countries without established national AI governance frameworks, intergovernmental and international initiatives present viable pathways for guiding the development of regulatory systems. For instance, the AU’s *Continental AI Strategy* aims to address governance challenges across the continent, where we found that 35 countries or territories have no AI-specific frameworks or legislation.^4^ In Latin America, 17 countries formalized their commitment to regional AI governance through the adoption of the *Cartagena Declaration on AI* in August, while the ASEAN *Guide on AI Governance and Ethics*, published in February 2024, provides a framework for cooperation and governance in Southeast Asia.^43,92^ While non-binding, these frameworks may help advance regional AI governance by promoting the adoption of AI-specific regulatory frameworks through standardized principles, common approaches, and coordinated mechanisms to address the ethical, legal, and socioeconomic implications of AI governance and deployment.

In contrast to non-binding intergovernmental frameworks, the Council of Europe’s *Framework Convention on AI*, adopted in May 2024, establishes the first legally binding international treaty on AI governance.^13^ The Convention requires that AI systems adhere to principles rooted in the protection of human rights, democracy, and the rule of law. To ensure compliance, the Conference of the Parties monitors adherence, provides recommendations, and facilitates collaboration through mechanisms such as public hearings. Member states are obligated to maintain transparency and accountability by documenting AI systems, ensuring accessible information for affected individuals, providing avenues to challenge AI-driven decisions, and establishing formal complaint mechanisms. As of December 2024, nine non-EU states with varying levels of progress in the adoption of horizontal AI regulation, including Andorra, Iceland, Israel, Moldova, Montenegro, Norway, San Marino, the UK, and the US, have signed the Convention. Notably, none of these nine countries have yet adopted their own horizontal binding frameworks for AI-specific governance. In this context, the Framework Convention serves as a promising solution to engage in collective governance, setting minimum standards while allowing flexibility for national adaptation. However, it remains to be determined whether such high-level international frameworks will be sufficiently implemented in legislation and enforced in individual countries.

In conclusion, the global landscape of AI governance is characterized by substantial disparities, marked by the uneven introduction of AI-specific regulatory frameworks and binding legislation, particularly between the Global North and South. Addressing these inequities necessitates international cooperation and the establishment of robust governance mechanisms. These mechanisms must prioritize inclusivity, equity, and adaptability to ensure the responsible deployment of AI technologies while promoting equitable access and opportunities across diverse socioeconomic contexts. However, while new AI-specific regulations may offer tailored approaches to emerging challenges, the absence of such legislation does not imply a lack of oversight for AI-based medical devices, as many are already governed under existing regulatory frameworks, with no evidence at present to suggest that updated guidance and standards would be any less effective in regulating AI-driven healthcare solutions.

## Data Availability

The generated dataset, including AI-specific frameworks and legislation for 197 countries and territories, will be made publicly available under the Creative Commons Attribution 4.0 International (CC-BY-4.0) license on figshare (https://figshare.com) upon acceptance of the publication. The digital object identifier will be provided upon acceptance of the publication.

## Acknowledgements

This work is supported by the European Union under the Horizon Europe Program as part of the project CYMEDSEC (101094218), ASSESS-DHT (101137347), and COMFORT (101079894). Views and opinions expressed are, however, those of the authors only and do not necessarily reflect those of the European Union. The European Union cannot be held responsible for them. The funding had no role in the study design, data collection and analysis, manuscript preparation, or decision to publish.

## Contributors

Conceptualization: Felix Busch, Stephen Gilbert, Esteban Ortiz-Prado, Keno K Bressem; Project administration: Felix Busch; Resources: Felix Busch, Stephen Gilbert, Esteban Ortiz-Prado, Keno K Bressem; Software: Felix Busch; Data curation: Felix Busch, Raym Geis, Yuan-Cheng Wang, Noor Al Khori, Israel K Kolawole, Warren Clements, Stephen Gilbert; Funding acquisition: Stephen Gilbert; Investigation: Felix Busch, Raym Geis, Yuan-Cheng Wang, Noor Al Khori, Israel K Kolawole, Warren Clements, Stephen Gilbert; Methodology: Felix Busch, Raym Geis, Yuan-Cheng Wang, Noor Al Khori, Israel K Kolawole, Warren Clements, Stephen Gilbert; Supervision: Felix Busch, Stephen Gilbert, Esteban Ortiz-Prado, Keno K Bressem; Validation: Felix Busch, Raym Geis, Yuan-Cheng Wang, Jakob Nikolas Kather, Noor Al Khori, Marcus R Makowski, Israel K Kolawole, Daniel Truhn, Warren Clements, Stephen Gilbert, Lisa C Adams, Esteban Ortiz-Prado, Keno K Bressem; Visualization: Felix Busch; Writing – original draft preparation: Felix Busch, Raym Geis, Yuan-Cheng Wang, Noor Al Khori, Israel K Kolawole, Warren Clements, Stephen Gilbert; Writing – review & editing: Felix Busch, Raym Geis, Yuan-Cheng Wang, Jakob Nikolas Kather, Noor Al Khori, Marcus R Makowski, Israel K Kolawole, Daniel Truhn, Warren Clements, Stephen Gilbert, Lisa C Adams, Esteban Ortiz-Prado, Keno K Bressem.

